# Design and Validation of a Compact Sensor for Peripheral Pulse Waveform Capture

**DOI:** 10.64898/2026.11.11.26367888

**Authors:** A. J. A. Naim, R. D. B. Ranaweera, D. Nandadeva, K. B. Divulwewa, A. Ratnayake, J. V. Wijayakulasooriya

**Affiliations:** Dept. of Electrical & Electronic Eng., University of Peradeniya, Peradeniya, Sri Lanka; Dept. of Physiology, Faculty of Medicine, Univ. of Peradeniya, Peradeniya, Sri Lanka; Cardiology and Cardiothoracic Unit, National Hospital Kandy, Kandy, Sri Lanka; Dept. of Anesthesiology & Critical Care, Faculty of Medicine, Univ. of Peradeniya, Peradeniya, Sri Lanka

**Keywords:** Peripheral pulse waveform, arterial tonometry, Hall effect sensor, non-invasive sensing, cardiovascular screening, pulse wave analysis

## Abstract

Peripheral pulse waveforms captured across different sites of the vascular tree provide valuable insights into cardiovascular health. This study presents a low-cost compact sensor designed to minimize intrusion at the measurement site while capturing the pulse waveform. The sensor performance was characterized and evaluated through both in-vitro benchtop experiments and a pilot in-vivo clinical study against invasive arterial pressure waveforms. The proposed sensor was able to capture pulse waveform morphology with reliable beat to beat tracking. Although offsets were present in the calculated features, fundamental temporal trends and cardiac cycle profiles were preserved. Thus, this work lays the foundation for a scalable hardware platform capable of multi-site pulse monitoring, enabling its integration with emerging computational frameworks for cardiovascular risk screening.

## I. Introduction

Non-invasive assessment of cardiovascular health is of growing importance due to the increasing prevalence of cardiovascular diseases [1]. In particular, low- and middle-income countries have limited access to early diagnostic care due to healthcare infrastructure constraints and the high cost of existing procedures. Therefore, it is important to have scalable, low-cost, and non-invasive screening tools to help clinicians refer patients for further tests at the community level. The peripheral pulse waveform serves as a rich physiological signature that reflects key hemodynamic parameters such as systemic vascular resistance, arterial stiffness, and aortic flow dynamics [2]–[4]. Numerous studies have established strong correlations between the pulse waveform and various vascular pathologies such as atherosclerosis, myocardial infarction, and stroke [5]–[7]. Leveraging the diagnostic potential of the peripheral pulse waveform thus offers a viable, non-intrusive pathway for early-stage cardiovascular screening in resource-limited settings.

Various physical sensing modalities have been presented in the literature to capture peripheral arterial pulse waveforms. Recent work has demonstrated the ability to measure the peripheral pulse waveform using bio-impedance [8]. While this method is immune to the skin tone based variations known to be present in PPG sensors, a major drawback is the requirement of subject specific calibration to account for the nonlinear effects introduced by anatomical differences and Body Mass Index. Alternatively, Hall-effect magnetic transducers have been introduced to measure digital artery pulse waveforms in previous work, though their application remains limited to the distal extremities [9], [10]. Adopting Hall-effect sensors in a clip type form factor for the acquisition of radial pulse waveforms has also been explored in previous work [11]. However, this arrangement cannot be readily scaled to non-planar peripheral sites such as the carotid and posterior tibial locations. As inverse hemodynamics and computational modeling methods emerge to map systemic cardiovascular dynamics, single-site pulse waveform analysis becomes increasingly restrictive [12], [13]. Fully leveraging these computational frameworks requires compact, non-intrusive sensor form factors capable of multi-site peripheral deployment without perturbing local vessel mechanics or altering blood flow dynamics. To address these challenges, this study focuses on the design, bench characterization, and pilot in-vivo feasibility evaluation of a compact Hall-effect sensor module suitable for multi-site peripheral arterial pulse capture.

## II. Methods and Materials

### A. Sensor Design

Fig. 1 shows the design of the pulse waveform sensor and its cross section. The sensor consists of a 3D printed enclosure housing a linear Hall effect sensor (AH49E, Diodes Inc.). An axially magnetized, cylindrical N52 Neodymium magnet (∅3 mm× 2 mm) is placed on a rubber membrane (AZDENT, Henan, China) of 0.18 mm thickness such that it stays vertically below the Hall sensor and attached to the base of the enclosure with a spacing of 7 mm. An arched cut extending 6 mm above the membrane along the longitudinal axis prevents the enclosure from compressing the artery, which would otherwise cause wave reflections at the contact points. A semi-circular bead centered on the rubber membrane concentrates contact directly over the artery, reducing the external pressure to capture the pulsations with high fidelity. This design allows it to be placed on the brachial, radial and tibial sites. The sensor is capable of measuring the pulsations at the carotid sites, although it requires an operator to manually place the sensor to capture the pulsations without compromising the breathing comfort of the subject. By utilizing standard off-the-shelf components this sensor is able to achieve a total Bill of Materials cost under $5.00, providing a significant cost advantage over commercial tonometers.

**Fig 1.**
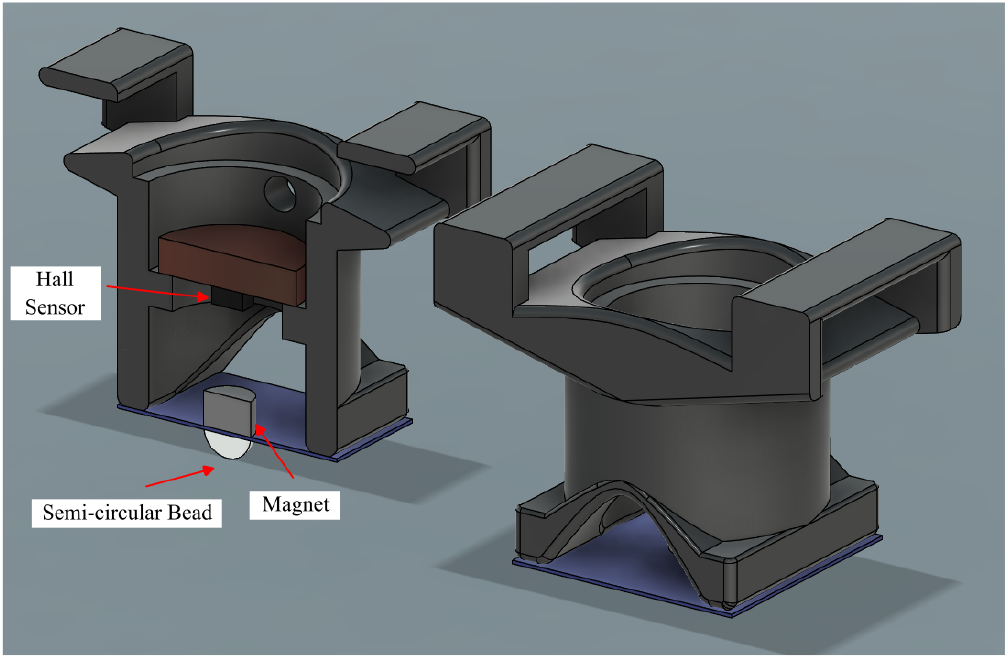
Design of the pulse waveform sensor and its cross section.

### B. Magnet Characterization

In order to characterize the magnets used in our sensor, the apparatus shown in Fig. 2 was designed to place the magnets relative to the transverse probe of a digital Gauss meter (Model TD8620-X, Tunkia Co., Ltd., Changsha, China). The magnets were centered on the probe’s active element and moved along the axis normal to its surface. Direct surface contact between the probe and magnet defined the zero-displacement reference point. Then the magnets were positioned from the probe in discrete steps of 0.3 mm up to a maximum distance of 12.3 mm, with step displacements verified using a precision dial caliper (RS Baty, RS Components, UK). Measurements were recorded at each discrete position during both the outward and inward movement of the magnet. The readings obtained for each set distance were averaged to reduce the errors due to any mechanical backlash in the setup. This procedure was repeated across *N* = 10 trials using distinct magnets in each run.

**Fig 2.**
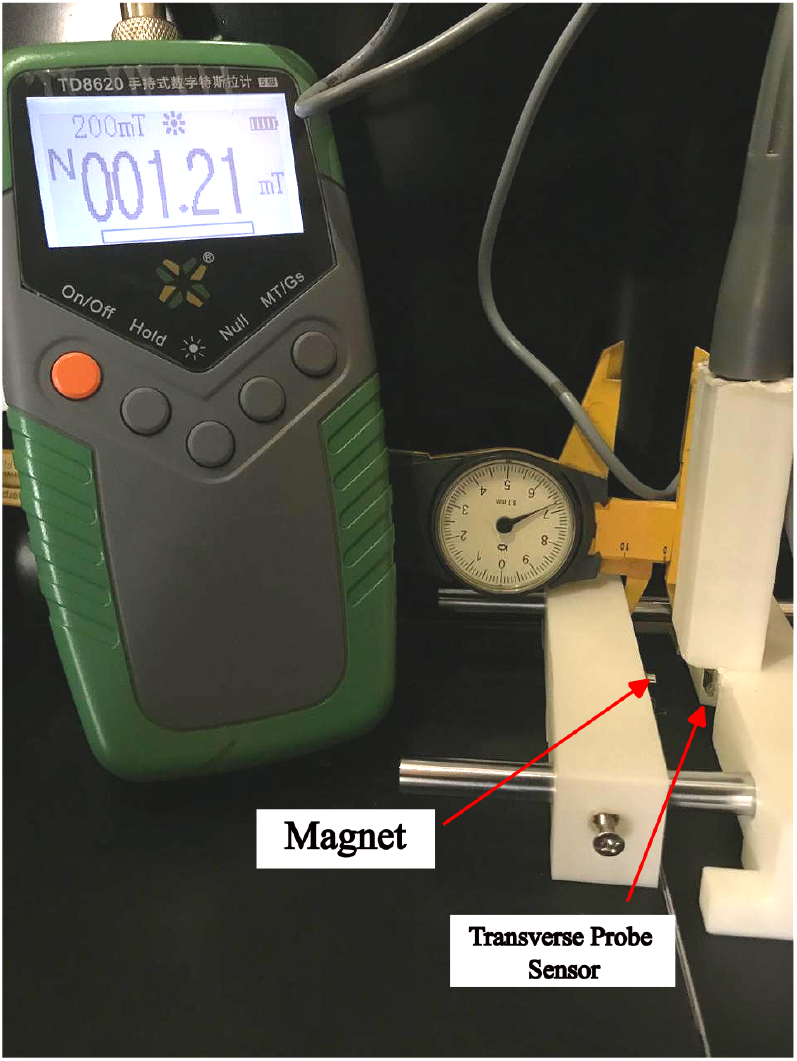
Experimental apparatus for magnet characterization relative to the Gauss meter transverse probe.

### C. Force vs Voltage Characterization

As the blood flow through the arteries generates a pulsatile force at the sensor interface, it is important to characterize the behavior of the rubber membrane as a function of the force applied. To perform this analysis, the setup shown in Fig. 3 was designed to secure the pulse waveform sensor and move it towards a digital balance with a resolution of 0.01g on a horizontally levelled platform. Prior to measurements the balance was calibrated against a known set of loads. A rigid cylindrical contact element was placed between the sensor and the balance to facilitate proper contact. Starting with initial surface contact, the sensor was moved downwards in steps of 0.5mm, up to a total displacement of 5mm. At each set distance the applied mass reading from the scale and the corresponding output voltage from the sensor were recorded simultaneously. This process was repeated across *N* = 5 distinct sensors. Measurements were recorded over the loading and unloading cycles for each sensor and averaged to minimize error due to any mechanical backlash in the setup. Mass readings were subsequently converted to applied force in Newtons.

**Fig 3.**
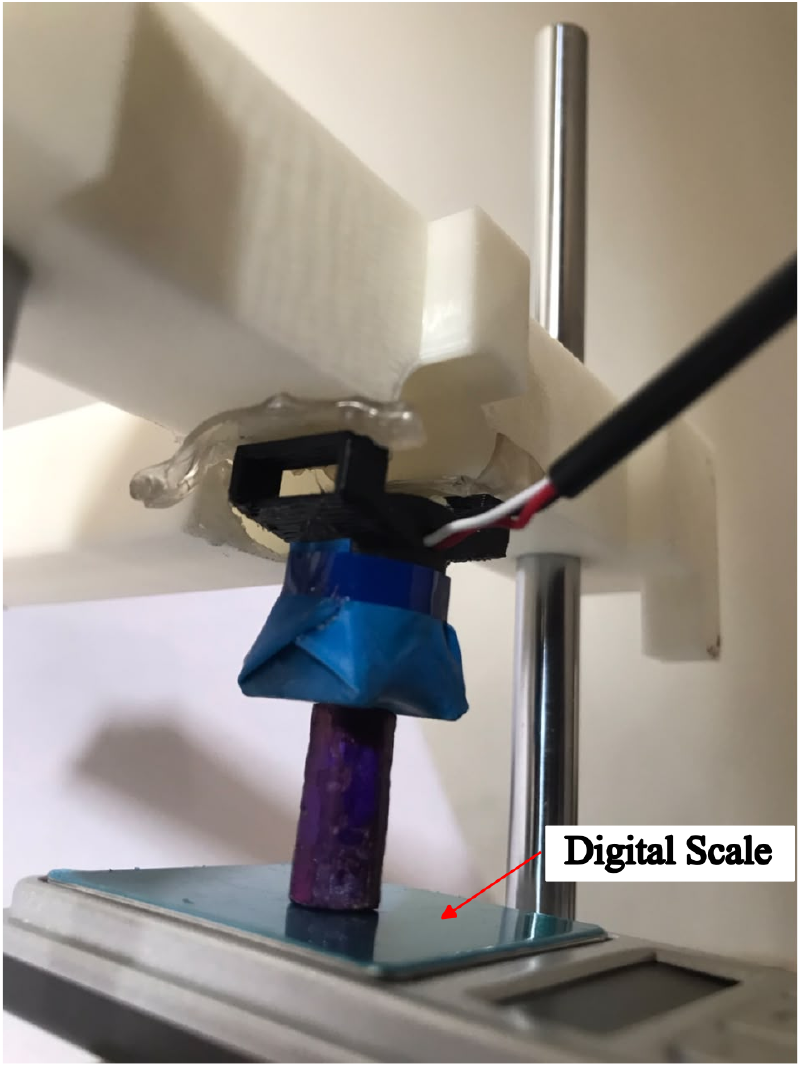
Experimental setup for force versus output voltage characterization.

### D. In vitro characterization

To characterize the dynamic response of the pulse waveform sensor, a peristaltic pump (KCM ODM, Kamoer Fluid Tech Co., Ltd.) was set up to circulate water through a flexible silicone model artery (inner diameter: 3.5 mm, wall thickness: 0.5 mm, length: 9 cm), as shown in Fig. 4. The pump was controlled to generate periodic pressure waves mimicking human arterial pressure profiles. Two arterial pressure sensors (MPX2300DT1, NXP Semiconductors) were connected at the proximal and distal ends of the model artery to continuously record invasive reference pressure waveforms. The pressure sensors were interfaced through instrumentation amplifiers (AD620, Analog Devices, Inc.) and calibrated using a 150 kΩ (0.1 %) precision resistor according to the datasheet. The pulse waveform sensor was placed directly at the center of the model artery to record the non-invasive waveforms generated as a result of the vessel displacement. All voltage measurements were obtained using a data acquisition module at a sampling rate of 1 kHz (USB-6009, National Instruments Corp.) Measurements were recorded for 30 seconds after the setup reached steady state operating conditions. This procedure was repeated for *N* = 5 trials using a distinct pulse waveform sensor in each run. The raw voltage signals were lowpass filtered with a cutoff of 15 Hz and downsampled to 125 Hz before analysis.

**Fig 4.**
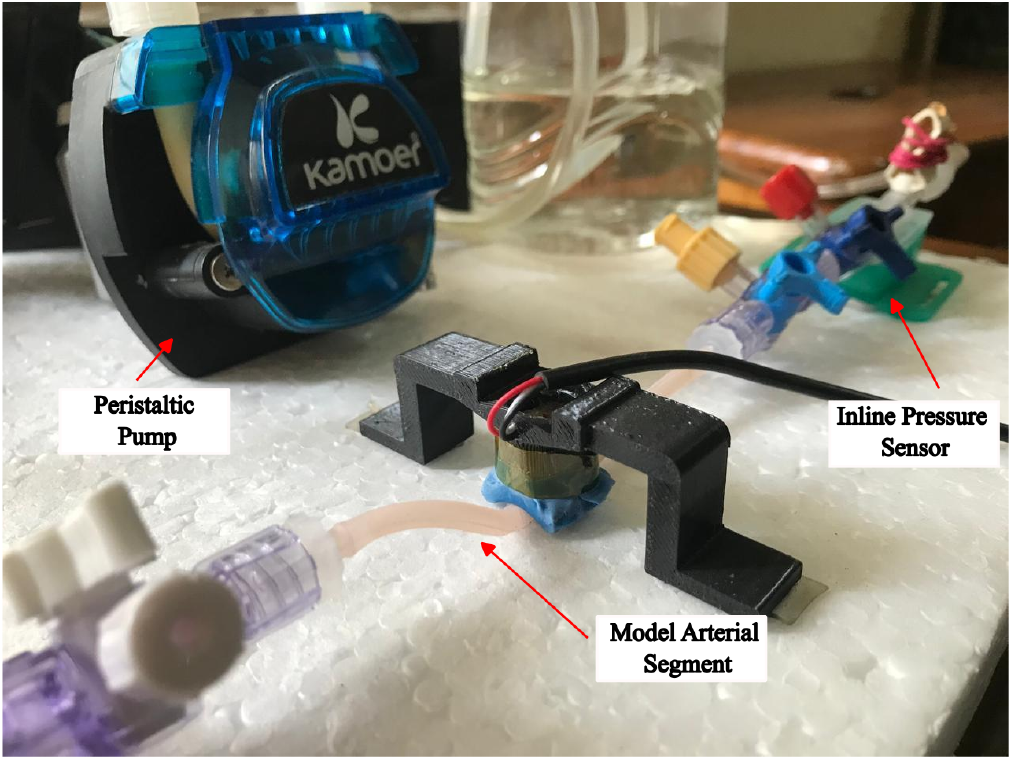
In-vitro characterization benchtop experimental configuration.

### E. Pilot In-vivo Clinical Evaluation

To evaluate the performance of the sensor against invasive arterial pressure waveforms, a pilot in-vivo feasibility study (*n* = 6) was conducted at the cardiothoracic ICU of the National Hospital Kandy under ethical approval (Ref no: NHK/ERC/22/2026) with written informed consent obtained from the patients. The sensor was placed at the ipsilateral brachial artery site while the patients were resting in the supine position with clinically prescribed in-line right radial catheterization for continuous blood pressure measurement. Invasive measurements were recorded using a Philips MX550 patient monitor via VSCapture software [15]. To synchronize the measurements between both devices, a blood pressure cuff was used to momentarily occlude brachial arterial blood flow. The signature of this response was later used to align the signals through cross correlation. The measurement system described in our previous work [14] was used to record the signals from the pulse waveform sensor at 1 kHz and later downsampled to 125 Hz to match that of the patient monitor signals. Waveforms were recorded for a duration of approximately 60 s. The signals were then low pass filtered with a 15 Hz zero-phase cutoff to remove any high frequency noise. Cubic spline interpolation was applied to the pulse onsets and subtracted to perform baseline drift correction and the signals were normalized. These signals were then segmented into individual cardiac cycles using the pulse onsets, to obtain 30 individual cycles for each participant. Pearson’s *r* was computed for each of the 30 invasive and non-invasive cycle pairs per participant and averaged. Overall agreement was reported as the mean of the six participant values. Five key pulse waveform features were extracted from the invasive and non-invasive signals using the PulseAnalyse toolbox [16]. Each feature was extracted from the ensemble-averaged beat of each participant and reported as mean*±* SD across participants. The paired difference was computed for each participant, and the mean difference was reported with a 95% confidence interval (CI) based on the *t*-distribution with 5 degrees of freedom.

All signal processing was performed offline in MATLAB (R2020b; The MathWorks, Inc., Natick, MA, USA)

## III. Results and Discussion

The spatial variation of the normal magnetic flux density (*B*) with relative distance is shown in Fig. 5. The average peak surface flux density was measured as *B*_max_ = 328.97*±* 3.30 mT. At the selected nominal separation of 7mm, the magnet produced a baseline flux density of 4.49*±*0.11 mT with a local small signal sensitivity of *S* = 1.95 mT/mm. However anatomical variations and changes in contact force can alter the operating distance between the sensor and magnet. Asliding window analysis (2.0 mm span) across *d* = 4.0 mm to 8.0 mm yielded a mean linearity of 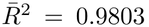. This demonstrates that small-signal pulse variations remain linear regardless of baseline placement offsets, eliminating the need for non-linear calibration.

**Fig 5.**
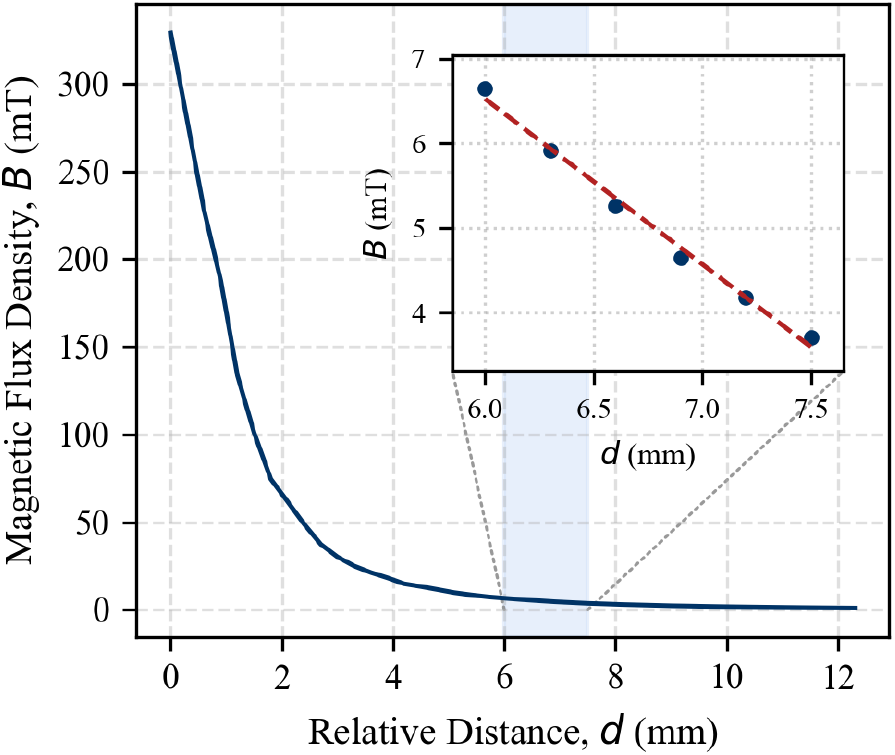
Spatial variation of the normal magnetic flux density (*B*) with relative distance, with inset showing the localized linear operating region.

The variation of sensor output voltage with applied interface force is shown in Fig. 6. This relationship exhibits a highly linear response with an *R*^2^ = 0.982 across the operational range. Defining 4.0 mm as the maximum baseline interface isplacement resulting from sensor contact with the arterial site, the measured force was 90.07*±*2.42 mN. Given the designed contact area of 2.2 cm^2^, this force corresponds to an interface pressure of 409.4 Pa. Compared to typical diastolic arterial pressures 9.33 kPa (70 mmHg) and 10.67 kPa (80 mmHg), the externally applied baseline pressure represents less than 4.5% of the arterial lower bound. This confirms that the sensor preload does not occlude the artery at the measurement site or perturb the local flow dynamics. Although Fig. 7 exhibits a non-linear relationship between output voltage and displacement, it is important to note that arterial pulsations occur in the sub-millimeter range [17]–[19]. Evaluating localized 2.0 mm sliding windows across 0–4.0 mm yields an average local linearity of *R*^2^ = 0.973*±*0.011, demonstrating that the localized small signal variations remain linear thereby preserving the pulse waveform morphology.

**Fig 6.**
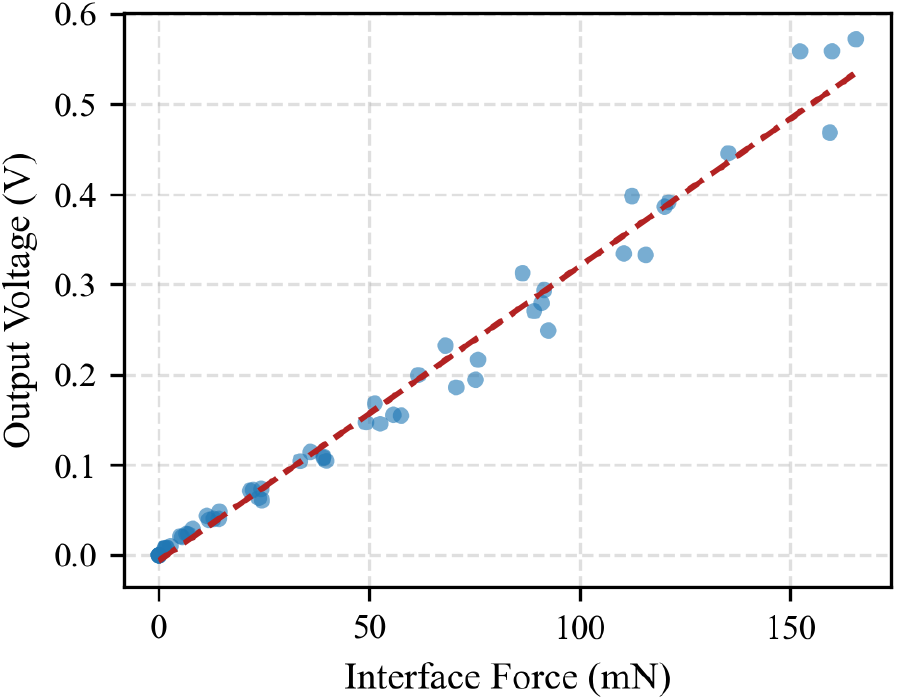
Variation of the force at the sensor interface with output voltage.

**Fig 7.**
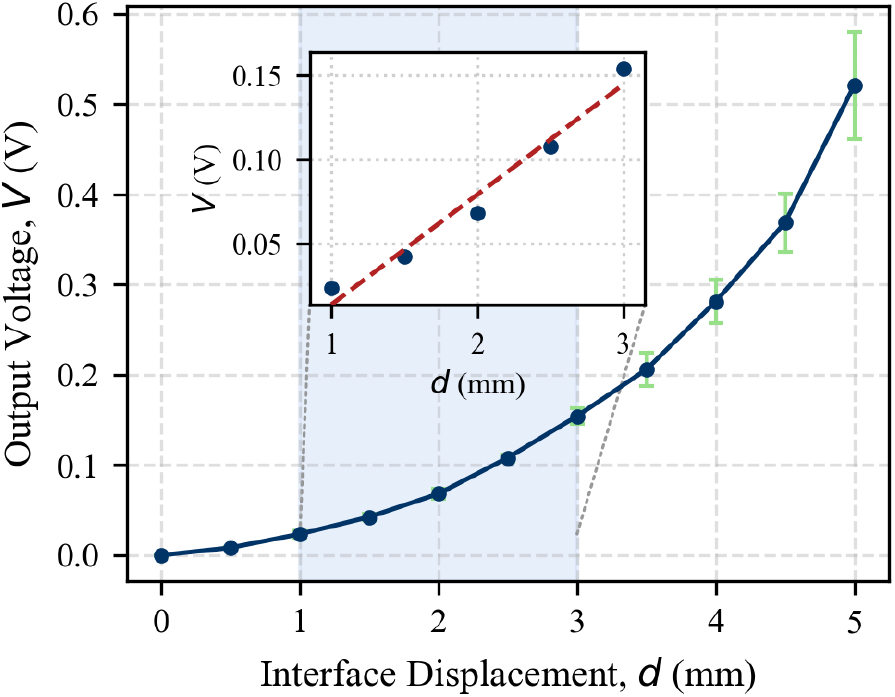
Sensor output voltage as a function of interface displacement, with inset highlighting the localized linear operating window.

During the in vitro characterization, the pulse waveform sensor displayed excellent dynamic tracking capability with respect to invasive pressures as shown in Fig. 8. The baseline conditions were set to yield proximal and distal mean pressures of 95 *±*3 mmHg and 80 *±*4 mmHg, with corresponding pulse pressures of 40 *±*4 mmHg and 34 *±*3 mmHg. Across all the trials, the pulse waveform sensor was capable of capturing the morphology of the pulse with beat-level correlations of *r* = 0.912*±*0.039 proximally and *r* = 0.935*±* 0.048 distally. The higher correlation with the distal sensor reading can be attributed to the natural damping of the vessel used to attenuate the ripples generated due to the pump motion. Following sensor placement, the mean pressure shifts remained minimal at +5*±*3 mmHg proximally and +3 *±*2 mmHg distally, while pulse pressure variations were limited to +4 *±*2 mmHg proximally and +2 *±*3 mmHg distally. This confirms that the sensor applied minimal disruption to native fluid dynamics. Although synthetic silicone arteries do not exactly replicate smooth muscle tone, these results confirm the sensor’s ability to measure the arterial waveforms with high fidelity without applying any significant obstruction to the flow.

**Fig 8.**
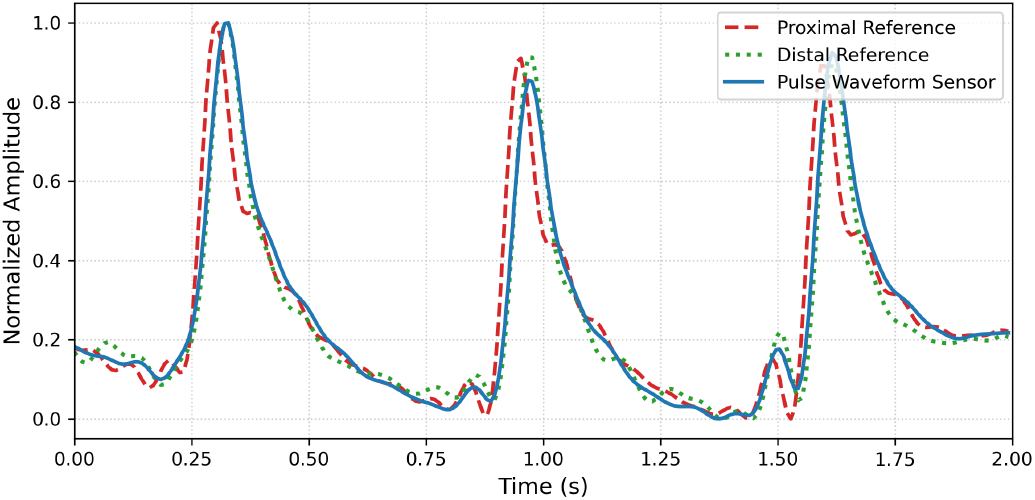
Dynamic waveform tracking comparing invasive reference pressure and non-invasive pulse waveform sensor output in the in-vitro model.

In comparison with invasive arterial pressure waveforms, the non-invasive pulse waveform signals demonstrated good morphological agreement. Overall linear correlation was high, yielding a mean per-subject Pearson correlation coefficient of *r* = 0.84 (range: 0.79 to 0.88 across subjects). A comparison of the invasive and non-invasive signals is shown in Fig. 9. While beat to beat trends are captured, it is clear that the non-invasive measurements have a wider systolic phase and an elevated diastolic phase compared to the invasive measurements, resulting in a higher area under the curve during systole (A1) and higher reflection index (RI, ratio of diastolic height to systolic height), respectively. The non-invasive measurements also displayed systematic offsets in key fiducial points such as the systolic peak and diastolic peak (secondary peak following the dicrotic notch), thereby introducing differences in the calculated Crest Time (time from pulse onset to systolic peak) and Δ*T* (time delay between systolic and diastolic peaks). Table I summarizes the features extracted using PulseAnalyse along with the mean difference between the signals and its 95% CI.

**Fig 9.**
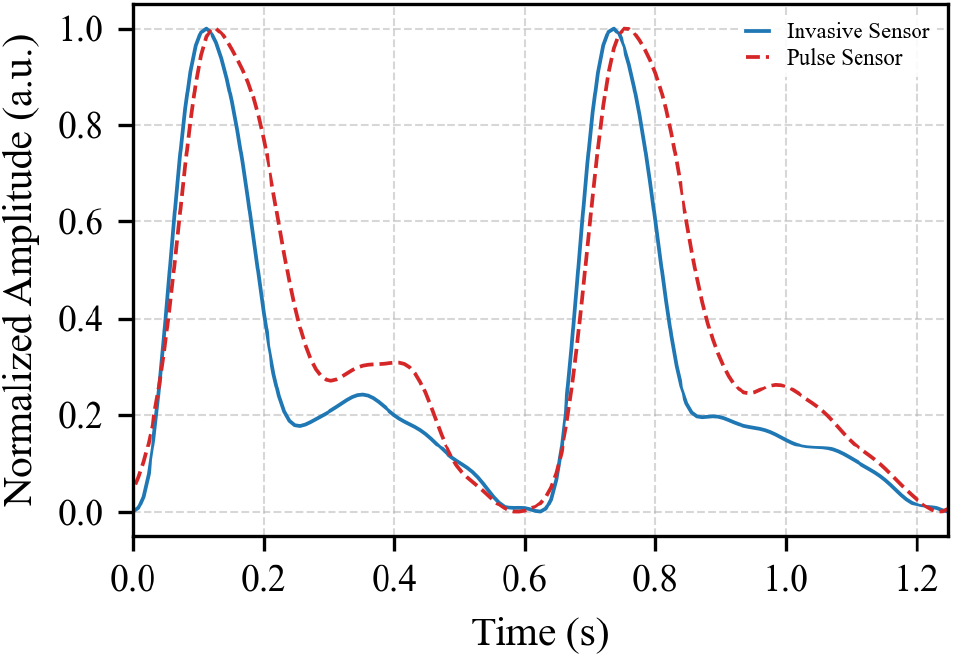
Comparison of in-vivo non-invasive brachial pulse waveforms and invasive radial arterial pressure waveforms.

It is important to note that the invasive sensor measures the internal pressure of the arteries whereas our non-invasive sensor measures the peripheral pulsations due to vessel wall displacement. Previous studies have established the non-linear relationship between the arterial pressure and arterial diameter [20]. Due to the viscoelastic nature of human arteries [21], [22], the arterial diameter can exhibit slightly different behavior between distension during the systolic phase and recoil during the diastolic phase within the same cardiac cycle. These factors may have contributed to the observations described above. Additionally, variations could have arisen due to the anatomical site difference between the radial and brachial locations as well as the compliance of the cutaneous tissue and subcutaneous fat present between the arterial vessel wall and the sensor interface.

**TABLE 1.**
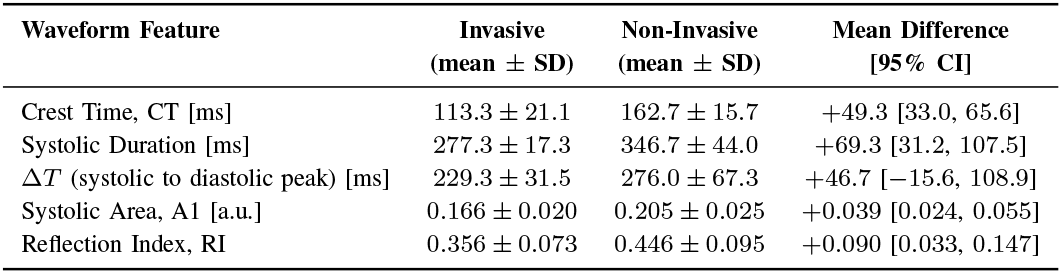
Comparison of Invasive and Non-invasive Arterial PulseWaveform Features.

## IV. Conclusion and Future Work

This study presented the design, characterization, and pilot clinical evaluation of a compact, non-intrusive sensor for peripheral arterial pulse waveform acquisition. In-vitro benchtop characterization confirmed that the sensor baseline placement force neither occludes the underlying vessel nor perturbs local fluid dynamics. Furthermore, in-vivo clinical evaluations demonstrated good morphological agreement between the non-invasive pulse measurements and invasive arterial pressure references. Differences were observed in features extracted from the non-invasive signal compared to the invasive reference, likely due to the different physical quantities measured by the two methods.

To address the differences present between invasive pressure and vessel wall displacement, future work will evaluate existing pressure-diameter models to compensate for the non-linear and viscoelastic behavior of arteries. Future research will also focus on the simultaneous deployment of the sensor platform across multiple peripheral sites, including the carotid, brachial, and posterior tibial arteries. This multi-site architecture will lay the foundation for non-invasive assessment of local and regional Pulse Wave Velocity, continuous cuffless blood pressure monitoring, and early cardiovascular risk screening.

## Data Availability

All data produced in the present study are available upon reasonable request to the authors

## ACKNOWLEDGMENT

This work was funded by grant MRG/2025/15 from the University Research Council, University of Peradeniya. The authors would like to express their sincere gratitude to the medical officers, nursing staff, and clinical personnel of the Cardiothoracic Unit at the National Hospital Kandy for their invaluable technical assistance, clinical support, and facilitation of patient data collection during the in-vivo validation study.

